# Substantia Nigra MRI markers are lower in Alzheimer’s disease and are linked to novelty and memory

**DOI:** 10.1101/2024.10.19.24315797

**Authors:** Friedrich Krohn, Mousumi Sarkar, Hartmut Schütze, Panagiotis Iliopoulos, Lucía Penalba-Sánchez, Dorothea Hämmerer, Renat Yakupov, Falk Lüsebrink, Annika Spottke, Anja Schneider, Nina Roy, Enise I. Incesoy, Michael Heneka, Ingo Kilimann, Luca Kleineidam, Stefan J Teipel, Frederic Brosseron, Doreen Goerss, Wenzel Glanz, Matthias Schmid, Ayda Rostamzadeh, Michael Wagner, Klaus Fliessbach, Frank Jessen, Emrah Düzel, Matthew Betts

## Abstract

Individuals with Alzheimer’s disease dementia (ADD) show Alzheimer’s disease (AD) pathology in and heterogeneous degeneration of the Substantia Nigra (SN) postmortem. However, it is unclear how the SN degeneration is related to cognitive dysfunction across the ADD continuum. In this study, using data from a prospective dementia study (DELCODE), we investigated if in-vivo SN MRI measures are lower in cases with clinically defined ADD than in healthy control subjects (HC) and associated with functional activity during the processing of novel visual stimuli and subsequent recognition memory.

161 DELCODE participants (69 years ±6 years, 88 men), including 79 Healthy Controls, 71 individuals with subjective cognitive impairment (SCD), 17 individuals with mild cognitive impairment (MCI), and 10 individuals with ADD, completed a scene novelty and encoding task and a 3T neuromelanin (NM)-sensitive MRI scan, from which in-vivo measures of SN MRI contrast and SN volume were calculated. For 71 individuals, CSF levels of phospoTau, total tau, and amyloid beta 42/40 ratio (Aß42/40) were available. All individuals completed a neuropsychological task battery from which a global cognitive score was calculated.

SN volume but not SN MRI contrast was lower in individuals with clinical ADD (One-way ANOVA: F(3,156)=4,13, N=160, p=0.0056, Tukey posthoc test: p=0.003) and SN MRI contrast and and volume were not associated with Aß42/40, ptau, and ttau CSF levels (all p<0.1). SN MRI contrast was positively associated with anterior hippocampal functional activity during the presentation of novel stimuli in individuals with Aß42/40 ratio levels below a pathological threshold of 0.08 (Aß42/40 positive, (p(FWE)=0.002, 157 voxels, n=28) but not in Aß42/40 negative individuals. Moreover, SN volume was positively associated with recognition memory (N=160, p=0.017, r^2^=0.11) and global cognition (p*<*0.0001, N=160, r^2^=0.221) across the ADD continuum.

Our study emphasizes the potential of using in-vivo SN MRI markers to understand the impact of AD pathology-independent SN degeneration on recognition memory and novelty processing dysfunction in ADD. Our results motivate future longitudinal studies exploring how SN volume and SN contrast change over time, how these differentially associate with cognitive decline and how the SN volume and SN contrast might be associated with other dopamine-dependent cognitive functions.

## 1 Introduction

The substantia nigra (SN), a central dopaminergic region, is involved in various functions, including movement initiation,^1^ executive function,^2^ working memory, ^3^ long-term memory ^4^ and novelty processin.^5^ Indeed, the SN and the surrounding Ventral Tegmental Area (VTA) cells show increased fMRI and cellular activity when individuals are presented with novel stimuli ^5–7^. Dopaminergic hippocampal novelty signals facilitate information encoding into long-term memory.^8–11^ Therefore, the SN might be linked to novelty-related memory formation.

Different SN MRI contrasts and SN volume have been assessed non-invasively in vivo using neuromelanin (NM)-sensitive MRI ^4,12–15^ owing to the SN’s high levels of NM, a dark iron-rich ^16^ that has been shown to correlate with SN MRI contrast.^12^ In older adults, SN MRI contrast and volume measures have been linked to memory and cognitive control ^4,17,18^. SN MRI measures are sensitive to volumetric ^15^ and SN MRI contrast ^19^ decrease and working memory decline^14^ in Parkinson’s disease.

While the role of the SN has been extensively studied in aging, its role in AD is underexplored. AD is a neurodegenerative disease characterized by the presence of pathological β amyloid and hyperphosphorylated tau.^20,21^ A meta-analysis showed lower dopamine levels and D1 and D2 receptor density,^22^ indicating that the dopaminergic system is affected in AD. Also, SN volume and dopaminergic functions are affected by AD: Post- mortem cases show that SN volumetric reduction in clinically defined dementia due to Alzheimer’s disease (ADD) is mild on average ^23–25^ but more pronounced than healthy aging^23^. Postmortem studies indicate that SN volume in ADD is highly variable across individuals—some show little to no degeneration.^26^ In contrast, others show comparable degeneration to individuals with Parkinson’s disease in a subset of individuals ^24,27^ and increasing volumetric cell loss with increasing AD dementia status.^27^Moreover, tau and amyloid pathology has been shown in SN cells in 60-91% of cases with ADD ^23,25,27–30^ and insoluble tau tangle levels are increased postmortem inside the SN at advanced stages of AD.^27^

Individuals with ADD show both behavioral ^31^and hippocampal activation ^32^ deficits during novelty processing and do not show enhanced memory for novel compared to familiar events. ^33,34^ While postmortem studies show that the SN is affected in ADD, to our knowledge, no studies have investigated how SN degeneration is related to novelty and long-term memory deficits in ADD. Here, to assess the role of the SN in deficits in dopaminergic dysfunctions in ADD, using data from the prospective DZNE – Longitudinal Cognitive Impairment and Dementia Study (DELCODE), we assess the relationship between in-vivo SN MRI measures, novelty processing, and recognition memory using a combination of NM-sensitive MRI and task fMRI in individuals spanning the ADD continuum. We also investigate how SN MRI contrast and volume are affected by AD pathology in a subset of individuals with available CSF biomarker levels. Finally, we investigate whether the relationship between SN volume and contrast, novelty processing, and memory is influenced by CSF biomarkers of AD pathology load.

## 2 Method

### 2.1 Subjects

Our dataset is a subset from the multicenter DELCODE study (Jessen et al., 2018) containing 160 subjects (72 men, 69±6 years) made up of 79 healthy controls (HC), 55 participants with subjective cognitive decline (SCD), 17 participants with mild cognitive impairment (MCI), and 10 individuals diagnosed with ADD (for participant statistics see table 1). All participants were older than 60, fluent in German, and provided informed consent. HC were recruited through local advertisements, while SCDs, MCIs, and ADDs were recruited through (self) referrals. All subjects performing within 1.5 SD on subtests of the CERAD (Consortium to Establish a Registry of Alzheimer’s disease) and not complaining about memory problems were considered healthy elderly individuals (HC). Subjects performing within the age- sex- and years of education-adjusted healthy range of 1.5 SD on CERAD but complaining of memory problems to physicians in memory clinics were regarded as individuals with SCD per recent guidelines.^35^ Subjects performing below 1.5 SD on the age- sex- and years of education-adjusted delayed recall CERAD task and complaining of memory problems to physicians in memory clinics and had intact daily functioning were classified as individuals with MCI per recent guidelines.^36^ Finally, subjects who fulfilled the clinical criteria for ADD according to current standards ^37^ were classified as individuals with ADD. Individuals performing below 18 points on the MMSE were excluded. For exclusion criteria and further DELCODE study details, see ^38^ The Institutional Review Boards of all participating study centers of the DZNE approved the study. All participants gave written informed consent before inclusion in the study. DELCODE is retrospectively registered at the German Clinical Trials Register (DRKS00007966) (04/05/2015). Data handling and quality control are reported elsewhere. ^38^

**Table 1:** Overview of the demographics, cognitive test results, and AD CSF pathology load in the sample analyzed in this work. Stars denote the P value of a two-sided t-test compared to HC: *=p*<*0.05, **=p*<*0.01, ***=p*<*0.0001

|  | HC | SCD | MCI | ADD |
| --- | --- | --- | --- | --- |
| **Whole dataset** |  |  |  |  |
| **N** | 79 | 53 | 18 | 10 |
| **Age** | 67.2 (5.1) | 70.5 (6.1)** | 71.72 (7)* | 72.1 *(6.6) |
| **Sex (M: F)** | 28:51 | 30:24 | 11:7 | 3:7 |
| **SN volume[mm<sup>3</sup>]** | 109.3 (25.6) | 114 (27.5) | 102.3 (27.3) | 83.18 (17)** |
| **Years of education** | 14.8 (2.67) | 14.95 (3.04) | 13.53 (2.92) | 12.8 (2.7) |
| **MMSE** | 29.35 (0.95) | 29.02 (1.11) | 27.82 (1.42)*** | 22.9 (2.92)*** |
| **TIV [ml]** | 1353.61<br>(205.53) | 1405.38<br>(223.31) | 1472 (259.05) | 1445.56 (170.08) |
| **Subset with available CSF data** |  |  |  |  |
| **N** | 30 | 20 | 14 | 7 |
| **CSF Ttau (pg/ml)** | 322.1 (116.3) | 346.3 (178.5) | 614.1 (291.1)** | 883.7 (502.2)*** |
| **CSF Ptau (pg/ml)** | 45.6 (13.9) | 48.4 | 72.3 (32.9)* | 106.6 (69.2)*** |
| **CSF Aβ42/40 ratio** | 0.1 (0.02) | 0.1 (0.002) | 0.006<br>(0.003)*** | 0.005 (0.02)*** |
| **CSF**<br>**Aβ42/Ptau181**<br>**ratio** | 18.5 (5.5) | 17.0 (4.4) | 10.3 (8.0)** | 5.8 (6.0)*** |

### 2.2 Structural MRI scans

MRI data for the present study were acquired with Siemens scanners (Verio, Skyra systems) at the study centers in Berlin, Bonn, Magdeburg, and Rostock. For the analyses reported here, T1-weighted MPRAGE [3D GRAPPA PAT 2, 1mm^3^ isotropic, 256 x 256 px, 192 slices, sagittal, 5min, repetition time (TR) 2500 ms, echo time (TE) 4.33 ms, inversion time (TI) 110 ms, flip angle (FA) 7°] images were used. Whole brain T1-weighted fast low-angle shot (FLASH) NM-sensitive MRI images were acquired using the following parameters: 0.75 x 0.75 x 0.75 mm^3^ voxel size, 320 x 320 x 192 matrix, 5.56ms echo time, 20ms repetition time, 23° flip angle, 130 Hz/pixel bandwidth, 7/ 8 partial Fourier, and 13:50 min scan duration, as previously reported.^39^

### 2.3 Novelty task

Participants first performed a previously published^40^ and adapted^41^ fMRI novelty (Fig. 1 A) and a recognition memory test. During the novelty part (Fig 1 A), inside the fMRI scanner, participants labeled two pre-familiarized images of scenes interleaved with 88 novel images of scenes as indoor or outdoor. Stimuli were shown as 8-bit gray-scale images scaled to 1250x750 pixel resolution and matched for luminance; the viewing horizontal half angle was 10.05° (’Presentation’ software by Neurobehavioral Systems Inc.). All participants underwent vision correction with MR-compatible goggles according to standard operating procedures. All sites used the same 30" MR-compatible LCD screen, matched for distance, luminance, color, and contrast across sites and the same response buttons. Stimuli were shown for 2500 ms each. fMRI images were recorded using the following parameters: 3.5mm isotropic resolution, isotropic, 64 x 64 px, 47 slices, oblique axial/AC-PC aligned, 9 min, TR 2580 ms, TE 30 ms, FA 80, 206 volumes. The novelty part took around 11 min. A recognition memory part was then performed in front of a PC 70 min after the fMRI task: Participants rated the 88 recently presented scenes and 40 additional images on a 5-scale familiarity scale, with 1 indicating they were sure they did not see the image, 3 indicating they are not sure if saw the image, and 5 indicating they were sure they previously saw the image.

**Figure 1.**
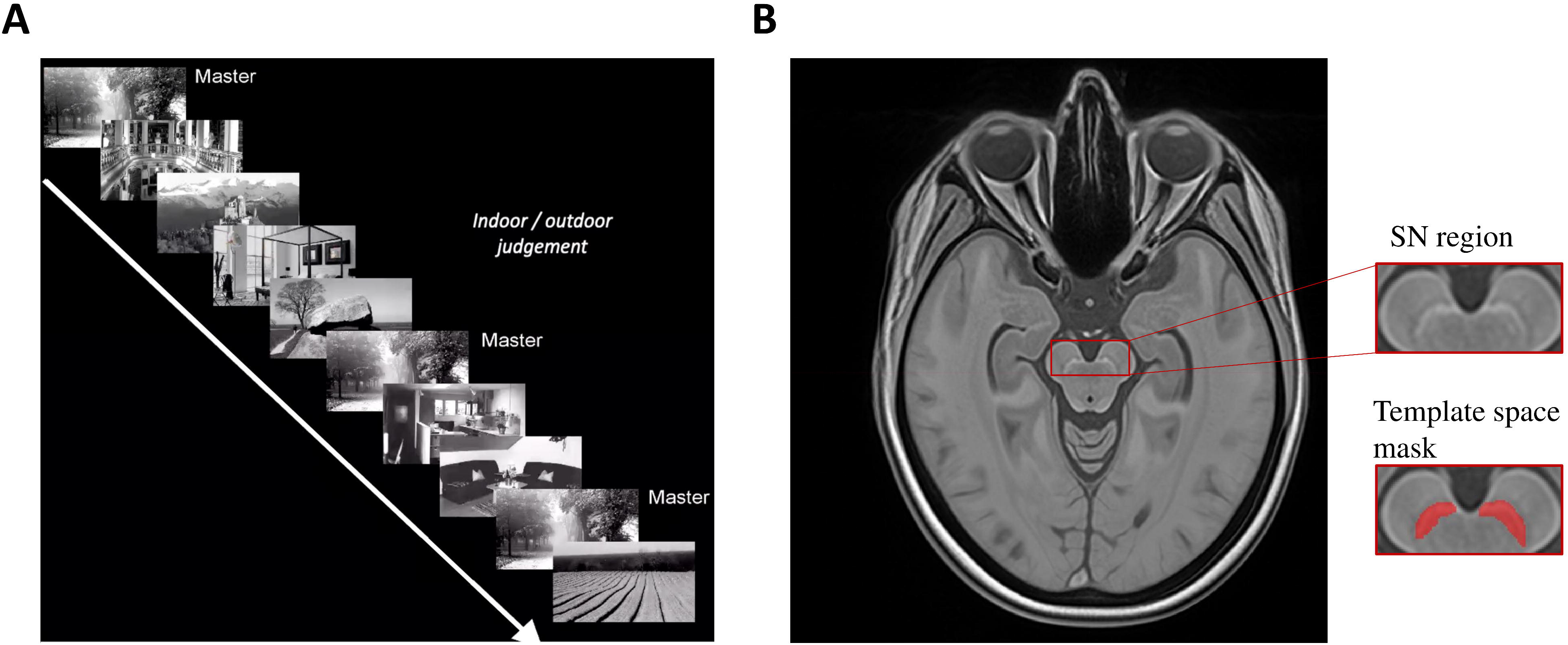
**fMRI task and SN template**: **(A)** Subjects were presented with 88 novel scenes interleaved with two pre-familiarized scenes, and they were instructed to label the images as inside or outside. 70 min later, they rated the 88 previously presented images and 44 novel images by their familiarity in front of a computer. **(B)** We generated a template image from 188 FLASH images using ANTs, from which the SN was segmented manually and morphed into subject space. **(C)** A pons mask (in yellow) was segmented in template space and morphed into native space for SN intensity normalization.

### 2.4 fMRI preprocessing

First-level general linear models were calculated after preprocessing (slice time correction, unwarping, realignment, and spatial smoothing (isotropic Gaussian kernel of FWHM 6 x 6 x 6mm) using Statistical Parametric Mapping (SPM), Version 12) in native space (including six motion regressors from the realignment process). We used a hemodynamic response function with a 128-s high-pass filter, no global scaling, and no serial correlations modeled. The novelty contrast was calculated as the difference between activation in response to novel and activation in response to familiar images.

### 2.5 Group-level analyses

Using the Advanced Normalization Tools toolbox, ^42^ a study-specific group template was calculated from T1-weighted MPRAGE images. Four rigid-then-affine iterations and six runs of a non-linear multiresolution routine ensured stable convergence (three resolutions, maximum of 90 iterations, template update step size of 0.1 mm). Individual native space novelty contrasts were warped to the group template space using the Advanced Normalization Tools toolbox function antsregistrationSyn. Voxelwise mass-univariate associations between structural SN MRI measures and hippocampal fMRI novelty activation were later performed in this space. Hippocampal masks were derived from the Desikan-Killiany atlas^43^ in FreeSurfer 6.0 (surfer.nmr.mgh.harvard.edu/). The FLASH images were upsampled to 0.375mm isotropic resolution using a MATLAB sinc function. Afterward, a study-specific template was generated from the FLASH images using the Advanced Normalization Tools toolbox (Fig 1B). In this template, the SN and a pons reference region (Fig 1C) were manually segmented using ITKSnap, and the segmented SN and pons were morphed back into the native space of all subjects. All masks were manually checked by expert raters (MS, MB). The SN MRI contrast was calculated as the ratio between the median intensity (int) inside the SN mask and the median int in the pons reference region as previously described^39^ using the following formula: SN volume was calculated using FSLmaths. We here use bilateral values as the sum between left and right. Manual and semiautomatic segmentations of 24 younger individuals and 43 individuals were generated by MB and MS as previously described^39^ and their volume and SN contrast were calculated. For this dataset, the dice scores of the volume were 0.66±0.15, the ICC for the volume was 0.04 and the ICC for the SN contrast was 0.79. The ICC was so low because, compared to the manual segmentations, the semiautomated segmentations tended to overestimate the SN shown by high sensitivity of 93±5% and a low specificity of 35±13%.

### 2.6 Cognitive Measures

Subjects were tested with extensive neuropsychological test batteries spanning verbal, memory, attentional, language, and visual domains.^44^ Please refer to^44^ for a detailed list of the applied batteries and tasks. Using confirmatory factor analysis in Mplus, five cognitive domain scores were derived from the task batteries: language, memory, executive function, visuospatial abilities, and working memory, and a global cognitive score was calculated as previously described from the five cognitive scores. ^44^

### 2.7 CSF measures

For 71 participants of our dataset, CSF levels of AD biomarkers Aβ42/40 ratio, p-tau-181, and t-tau levels were measured. In adherence with recent guidelines in the ATN framework,^21^ we thus estimated the Aß42/40 ratio, tau pathology (p-tau-181), and neurodegeneration (total tau). The CSF levels were obtained centrally in Bonn using commercially available kits according to vendor specifications: V-PLEX Ab Peptide Panel 1 (6E10) Kit (K15200E) and V-PLEX Human Total Tau Kit (K151LAE) (Meso, MD, USA) and Innotest Phospho- Tau(181P) (81581; Fujirebio Germany GmbH, Hannover, Germany) as described previously.^38^ We binarized the subjects into pathology positive/negative (i.e., Aβ42/40 ratio smaller than 0.08 as Aβ42/40 positive, subjects with total tau levels greater than 510.9 pg/ml as tau positive, and subjects with levels greater than 73.65 pg/ml as phosphotau 181 positive) and vice versa according to recent guidelines.^45^

### 2.8 Statistical analyses

Analyses were performed in Matlab 2022a (Natick, Massachusetts). We used linear models to investigate the relationship between SN MRI markers, CSF biomarker level, and recognition memory. Using an ANCOVA, we also assessed the difference in SN volume and SN MRI contrast between diagnostic groups. We used the Statistical Parametric Mapping Toolbox 12 Revision 7771 (SPM) to calculate mass-univariate models to assess the relationship between SN MRI contrast and volume and hippocampal novelty contrast, applying family-wise error (FWE) cluster-level correction for multiple comparisons. We limited our voxelwise analysis to a region defined by a hippocampal mask. In all analyses, we used scanner site, age, total intracranial volume (TIV), years of education, and sex as covariates. Linear associations were plotted using the Gramm toolbox for Matlab^46^, and group comparisons were drawn using the Raincloud toolbox.^47^ We calculated Hedge’s g,^48^ which is similar to Cohen’s d but insensitive to group size differences, to calculate the effect sizes of the diagnostic group differences.

## 3 Results

A significant difference in SN volume between diagnostic groups was observed (ANCOVA; F(3,156)=4,13, N=160,p=0.001, Figure 2A). A subsequent Tukey-post-hoc test revealed that SN volume was lower in individuals with ADD compared to HC (p=0.003, 23% average volume difference) and compared to individuals with SCD (p=0.013, 28% average volume difference). SN volume of each HC (p=0.7) and SCD (p=0.36) did not differ from MCI SN volume. No significant difference in SN MRI contrast between diagnostic groups was observed (F(3,156)=0.23, N=160,p=0.89, Fig. 2B). We did not find an association between SN MRI contrast and SN volume (p=0.11, r^2^=0.07, n=160), indicating that the MRI measures are statistically independent. A one-way ANOVA revealed that SN volume (F(3,156)=3.05, p=0.03, N=160) and SN MRI contrast differed between scanner sites (F(3,156) = 51.57, p*<*0.001, N=160). Two-sided t-tests between the left and right SN volume and SN MRI contrast revealed a significantly higher SN volume (p*<*0.001) and significantly higher SN MRI contrast (p=0.0092) in the right compared to the left SN hemispheres. All tested associations are listed in supp. Table 1.

**Figure 2.**
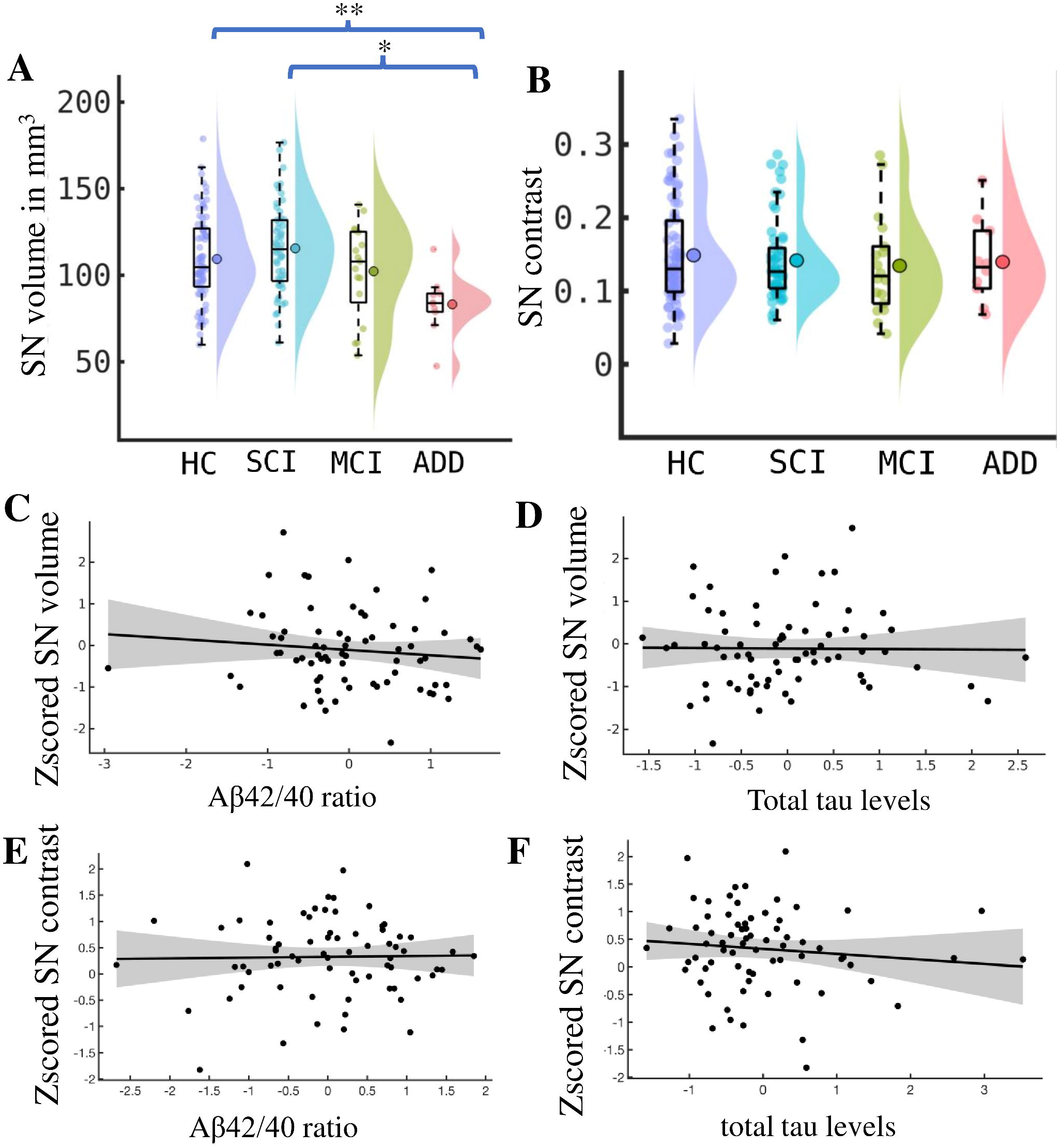
**SN volume is lower in ADD but is not associated with AD pathology**: **(A)** An ANCOVA between SN volume and diagnostic groups and a subsequent posthoc tests revealed a significantly lower SN volume in individuals with ADD compared to HC and SCD (F(3,156)=4,13, N=160,p=0.0056). **(B)** An ANCOVA between SN MRI contrast and diagnostic groups revealed no significant difference (F(3,156)=0.23, N=160,p=0.89). **(C)** No significant association between SN MRI measures and Aβ42/40 ratio (C, E) or total tau (D, F) was observed (n=71).

### 3.1 SN volume is reduced in ADD but is not associated with CSF Aß42/40 and tau levels

We performed a one-way ANOVA comparing diagnostic group differences in SN volume (F(3,156)=4,13, N=160, p=0.0056, Fig. 2A). A subsequent Tukey-post-hoc test revealed a lower SN volume in individuals with ADD compared to HC (p=0.003, Hedges’ g=1.056) and lower SN volume in individuals with ADD compared to individuals with SCD (p=0.013, Hedges’ g=1.16). There were no statistical differences in SN volume between HC and MCI (all p>0.05). The lack of statistical difference between HC and MCI indicates that SN volume decreases late into the disease progression. The significant effect size of g>1^48^ for group differences between HC and ADD and the non-significant association between HC and MCI indicate a sharp decrease in SN volume with clinical ADD onset. A one-way ANOVA comparing SN MRI contrast between diagnostic groups revealed no significant difference between groups (F(3,156)=0.23, N=160, p=0.89, Fig. 2B). Overall, these results indicate that the SN shrinks but maintains a stable level of neuromelanin in individuals with ADD.

However, we did not find any association between SN volume and SN volume and the CSF biomarker levels (all p>0.1, Fig. 2C-E).

### 3.2 Higher SN MRI contrast predicts higher voxel-wise hippocampal novelty contrast in Aβ42/40 positive individuals

To determine if SN MRI measures are related to changes in hippocampal activation during the presentation of novel stimuli, we calculated mass-univariate associations between SN volume, SN MRI contrast, and hippocampal novelty activation while correcting for site, age, sex, TIV, and years of education. These associations did not reveal any significant family-wise corrected- clusters. However, in Aβ42/40 positive individuals, SN MRI contrast was positively associated with the novelty contrast inside the hippocampus (p(FWE)=0.002, 157 voxels, n=28, Table 2). These results are depicted in the horizontal (Fig. 3A) and sagittal (Fig. 3B) plane. We did not find this association in Aβ42/40 negative individuals.

**Figure 3:**
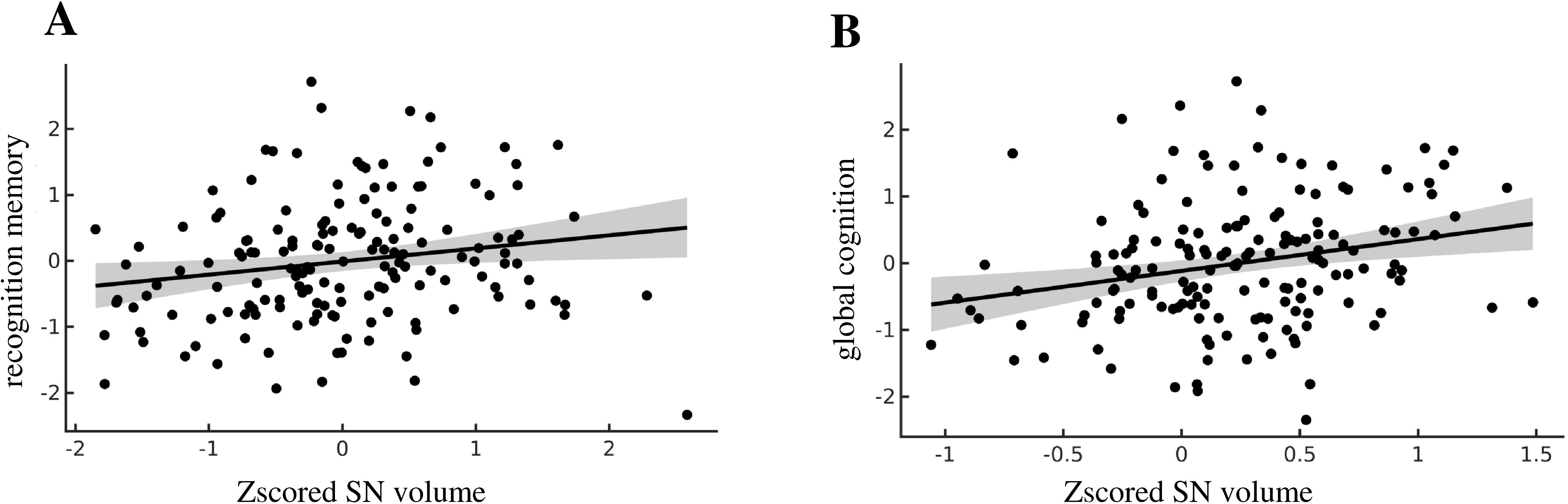
**Higher SN MRI contrast predicts higher hippocampal novelty contrast in Aß42/40 – positive individuals**: **(A)** Using mass univariate linear models, we identified an association between SN MRI contrast and a cluster in the anterior right hippocampus. Activation is zoomed in on the hippocampus in a 40x40 voxel window. **(B)** The same hippocampal cluster as in **(A)** is shown in the sagittal plane. Note that ‘T’ denotes each voxel’s T value statistics.

**Figure 4:**
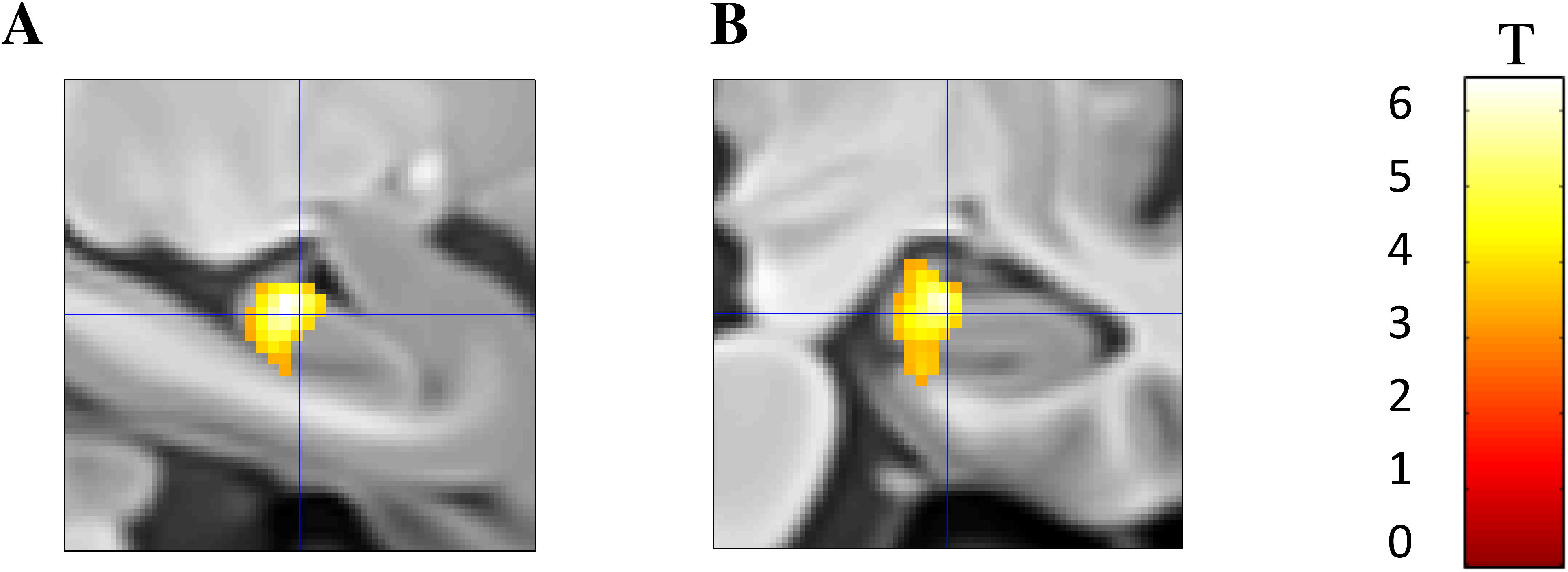
Linear regression models regressing SN volume and each recognition memory and global cognition were significant: **(A)** SN volume predicted better recognition memory (N=160, p=0.017, r2=0.11). **(B)** Higher SN volume predicted better global cognition. p=0001, N=160, r2=0.221)

**Table 2:** SN MRI contrast correlates with novelty activation in a cluster in the right anterior hippocampus independent of diagnostic group and hippocampal volume

| Cluster size | Cluster $p_{\text{FWE}}$ | Peak T | Template x,y,z (mm) | Brain structures | Additional covariates |
| --- | --- | --- | --- | --- | --- |
| 157 | 0.008 | 6.99 | 22 2 -18 | Right anterior hippocampus | none |
| 89 | 0.027 | 7.27 | 22 2 -17 | Right anterior hippocampus | hippocampal volume |

|  |  |  |  |  |  |
| --- | --- | --- | --- | --- | --- |
| 152 | 0.009 | 6.78 | 22 2 -18 | Right anterior<br>hippocampus | diagnostic<br>group |

The association between hippocampal novelty contrast and SN contrast in Aβ42/40 positive individuals was still observed after correcting for hippocampal volume (p(FWE)=0.027, 89 voxels, Table 2) and diagnostic group (p(FWE)=0.0.009, 152 voxels, Table 2), indicating that this finding was independent of hippocampal atrophy. This association was also observed in all individuals with known CSF biomarker status (p(FWE)=0.018, 181 voxels, N=71, Supplementary Table. 2)

### 3.3 Higher SN volume predicts higher recognition memory and a higher global cognitive score

A one-way ANOVA investigating diagnostic group differences in recognition memory (F(3,158)=19.5,p<0.001, N=160) and Tukey post hoc tests revealed significantly lower recognition memory in ADD and MCI subjects compared to HC and SCD (all p<0.001, suppl. table S2).

As SN volume and recognition memory were decreased in individuals with ADD, we corrected for clinical group to assess for associations between SN MRI measures and recognition memory. We found a positive association between SN volume and recognition memory (N=160, p=0.017, r^2^=0.11) across groups. This finding remained significant after correcting for hippocampal volume (p=0.002, r^2^=0.162) and diagnostic group (p=0.023, r^2^=0.231), indicating this effect was independent of AD-driven degeneration.

To determine if the SN volume was related to overall cognitive function, we performed an additional analysis associating SN volume and contrast with a global cognitive score aggregated across five different cognitive domains (visuospatial abilities, memory, working memory, language, and executive function). SN volume was positively associated with the global cognitive score (p<0.0001, N=160, r^2^=0.221, Fig. 3B). This association held after correcting for hippocampal volume (p<0.0001, N=160, r^2^=0.175) and diagnostic group (p=0.002, N=160, r^2^=0.56) indicating the association was not driven by AD. In addition to finding an association between SN volume and global cognition, we also found weak but significant correlations between SN volume and latent factors related to visual: p=0.007, r^2^=0.1, working memory: p=0.003, r^2^=0.125, language: p<0.001, r^2^=0.133 and executive function: p<0.001, r^2^=0.136, suppl. Fig. 1). These results also remained significant after correcting for hippocampal volume (Visual: p=0.03 r²=0.1, working memory: p=0.003 r²=0.12, language: p=0.001 r²=0.12735, executive function: p=0.001, r^2^=0.131). These results indicate that higher SN volume was associated with recognition memory and cognitive ability across multiple domains and are independent of hippocampal atrophy in individuals across the ADD continuum.

## 4 Discussion

Using in vivo structural MRI measures of the SN (SN volume and MRI contrast), we assessed how SN the in ADD is affected in ADD and how the SN is related to hippocampal activity during novelty, subsequent recognition memory, and general cognition. In a subset of individuals with CSF biomarker data, we also assessed how SN degeneration is related to AD pathology. Our analyses revealed that SN MRI contrast and SN volume are statistically independent measures and that SN volume is lower in individuals with ADD compared to HC and individuals with SCD. However, SN volume did not differ between HC and MCI and was not associated with CSF biomarkers of AD pathology. While SN volume was associated with poorer recognition memory, SN MRI contrast was associated with hippocampal novelty in Aß42/40 positive individuals but not in Aß42/40 negative individuals. Finally, we found that SN volume was associated with a decline in recognition memory and global cognition in ADD but not with CSF measures of AD pathology.

We found no association between SN MRI contrast and SN volume. Previous studies have employed both SN MRI contrast and SN volume measures but did not clarify the relationship between them.^49–52^ Our results indicate that they are statistically independent in the sample tested. Postmortem literature suggests that SN MRI contrast correlates with the concentration of NM-rich cells in the SN,^12^ and high SN MRI contrast spatially overlaps with clusters of cells producing tyrosine hydroxylase (TH).^53^ However, other authors suggest that the MRI contrast might also be driven by water concentration,^54^ indicating that the mechanisms underlying SN MRI contrast are still debated and not entirely understood. Several additional factors such as MRI contrast from blood vessels,^55^ astrocytes,^56^ myelinated axons,^57^ and non-dopaminergic neurons^58^ have been shown inside the SN and might also contribute to SN MRI contrast. Future studies are needed to investigate how SN-MRI contrast and SN volume are related to dopaminergic function and whether they can be used as statistically and functionally distinct indicators.

Our study revealed MRI-derived SN volume was 25% lower in ADD compared to HC. This decrease aligns with previous postmortem studies reporting varying degrees of SN volume reduction between a 10% and a 40% lower SN volume. ^24,59,60^ Also this study is the first to our knowledge, to report a reduction in SN volume in vivo. One study found no difference in SN volume^26^. Moreover, we found no SN volume decrease in MCI or SCD compared to HC, indicating that SN volume decreases occur at a later stage of ADD. These results indicate SN volume might be a sensitive in-vivo MRI marker to detect potential dopaminergic deficits in ADD. They pave the way for future longitudinal studies to assess how SN volume might be linked to changes in dopamine-dependent function across time. Interestingly, studies assessing group differences in NM-sensitive MRI contrast using TSE MRI sequences^61,62^ also found no significant decrease in SN MRI contrast in ADD, in line with our results presented here.

In a subset of individuals with CSF biomarker status, we found no significant association between either SN MRI contrast or SN volume with AD pathology. Several postmortem studies also showed no association between SN cell loss and tau or amyloid pathology.^25,28^ While one study^27^ found an increase in tau in the SN together with increased SN degeneration in AD, they were not directly associated. However, we are the first to show this extends to CSF levels of AD pathology. Collectively, these findings indicate that SN degeneration may not be linked to the extent of tau or amyloid pathology. Previous reports show alpha-synuclein is related to SN degeneration in ADD. In one study alpha-synuclein was moderately associated with SN loss in ADD.^28^ Alpha-synuclein is toxic to the SN pars compacta,^63^ notably in Parkinson’s disease.^64^ Additionally, elevated alpha-synuclein levels have consistently been demonstrated in individuals with MCI and ADD.^27,65–68^ Therefore, the SN degeneration observed in our study may be related to elevated alpha-synuclein levels. While alpha-synuclein was not assessed in DELCODE, it will be important in future studies to investigate to what extent alpha-synuclein pathology is related to SN degeneration in ADD. The results presented here indicate amyloid and tau-independent mechanisms may contribute to SN degeneration in ADD.

Previous studies revealed an inverted U-shaped association between Aß42/40 CSF levels and hippocampal activity during the novelty recognition task analyzed here,^32^ indicating that Aß42/40-positive individuals have lower hippocampal activity in later clinical stages (MCI, dementia). Additionally, CSF tau levels are only associated with hippocampal novelty activation in Aß42/40-positive individuals.^69^ We show here that SN MRI contrast is only associated with hippocampal novelty contrast in Aß42/40 positive individuals. Should higher SN contrast predict higher dopaminergic output,^12^ this would indicate that in individuals with decreased baseline hippocampal novelty activity, higher SN dopaminergic release might act as a compensatory mechanism to restore hippocampal novelty activation in individuals with high levels of amyloid. Alternatively, a decrease in SN volume and a decreased baseline hippocampal novelty activity are sufficient to decrease hippocampal novelty activity significantly. Higher SN MRI contrast,^4^ higher SN/VTA MRI activity,^7,70–75^ and higher cellular SN activity ^5^ predict better long-term memory. We extend these previous findings by showing that SN volume is related to memory decline in individuals spanning the AD continuum. We also expand previous findings showing that the SN is linked to broader cognitive functions such as cognitive control^2,17^ by showing that it is related to several factor scores spanning five cognitive domains and global cognition. Future studies should assess how SN volume is related to other dopaminergic functions such as working memory^76^ and reward.^77^

It is important to note several limitations on interpretation of these findings. The number of individuals with ADD was low, thus the results presented here need to be replicated in larger samples. The contribution of VTA degeneration, another source of dopamine in the brain, in ADD,^78^ to novelty and memory deficits remain is to be investigated. Our SN segmentations were performed semi-automatically, possibly introducing bias and an SN overestimation. Finally, our study was cross-sectional, longitudinal follow-up data, once available, will enable us to explore how SN degeneration is related to cognitive function over time.

Future studies should attempt to replicate our results in larger cohorts of individuals with ADD with AD pathology CSF levels. They should also assess how SN contrast and SN volume are structurally and functionally related and to what extent they serve as independent in-vivo SN MRI markers. Further investigation is required toassess whether SN volume, as previously observed for SN MRI contrast, is related to other dopamine-dependent functions. Finally, further research is required to systematically assess the exact mechanisms underlying SN degeneration in ADD, e.g. with respect to the contribution of alpha-synuclein.

In conclusion, we show in-vivo SN markers are differentially related to hippocampal activity during novelty and recognition memory decline in ADD. SN degeneration is independent of CSF biomarkers of AD pathology, opening avenues for longitudinal assessments of tau- and Aß42/40 independent mechanisms of SN degeneration in ADD.

## Data availability

Data, study protocol, and biomaterials can be shared with partners based on individual data and biomaterial transfer agreements.

## Data Availability

All data produced in the present study are available upon reasonable request to the authors

## Acknowledgments

The authors would like to thank all study participants for their invaluable contributions.

## Funding

The study was funded by the German Center for Neurodegenerative Diseases (Deutsches Zentrum für Neurodegenerative Erkrankungen; DZNE), reference number BN012.

F.K is supported by the German Federal Ministry of Education and Research (BMBF, funding code 01ED2102B) under the aegis of the EU Joint Programme – Neurodegenerative Disease Research (JPND). He is paid from a stipend awarded by the medical faculty in Magdeburg.

E.D. and M.B. are supported by Deutsche Forschungsgemeinschaft (DFG, German Research Foundation)—Project-ID 425899996—SFB 1436

## Competing interests

The authors declare no conflicts of interest.

## Funding

The study was funded by the German Center for Neurodegenerative Diseases Deutsches Zentrum für Neurodegenerative Erkrankungen (DZNE)], reference number BN012, the European Union’s Horizon 2020 Research and Innovation Programme under Grants 785907 (HBP SGA2) and 945539 (HBP SGA3), and the Deutsche Forschungsgemeinschaft (DFG, German Research Foundation)—Project-ID 425899996 – SFB 1436.

## Abbreviations

SN= Substantia Nigra, HC=Healthy Control, SCD=Subject cognitive decline, MCI=Mild cognitive impairment, ADD=clinically defined dementia due to Alzheimer’s disease

---

**Figure S1:**
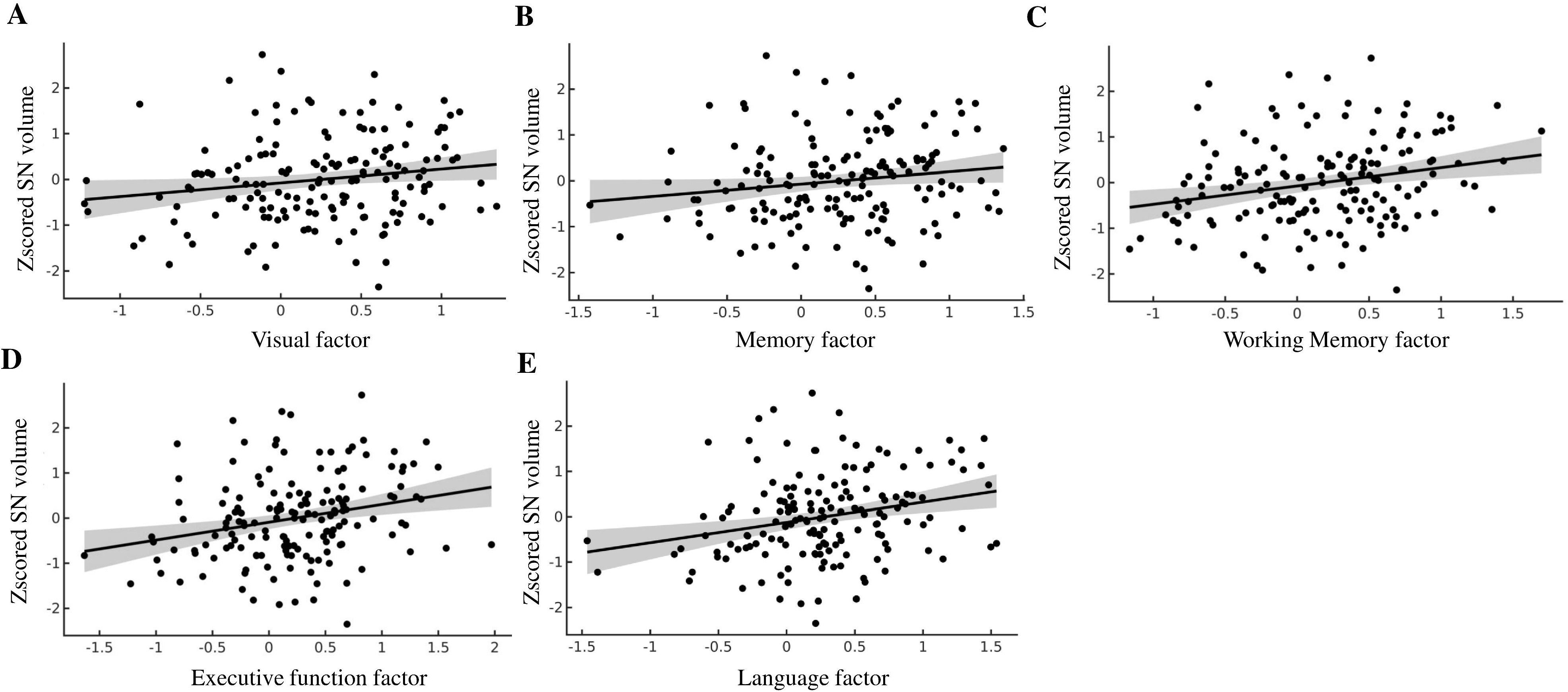
Higher SN volume predicts higher values of all factor scores after correcting for site, years of education, sex, and age: **(A)** SN volume correlates with visuospatial abilities (p=0.0007,n=160, *r*^2^=0.281) **(B)** SN volume correlates with memory (p=0.007,n=160, *r*^2^=0.1) **(C)** SN volume correlates with working memory (p=0.007,n=160,*r*^2^=0.328) **(D)** SN volume correlates with executive function (p*<*0.001,n=160, *r*^2^=0.387) **(E)** SN volume correlates with language (p*<*0.001,n=160, *r*^2^=0.397

**Table S1:**
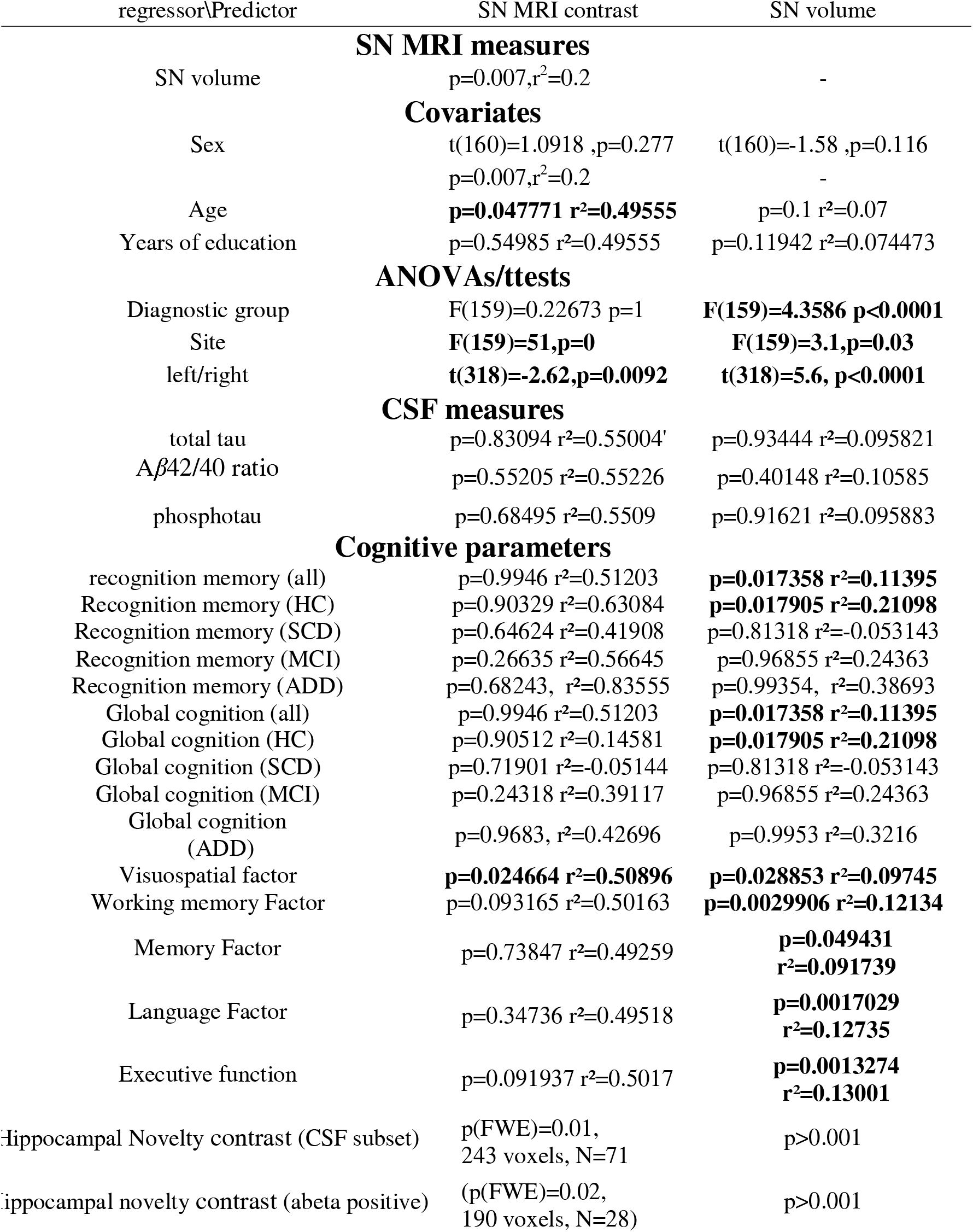
Overview of all tested linear correlations with years of education, age, sex, and scanner site as covariates, r^2^ indicates Pearson correlation coefficient. Significant correlations are highlighted in bold.

**Table S2:**
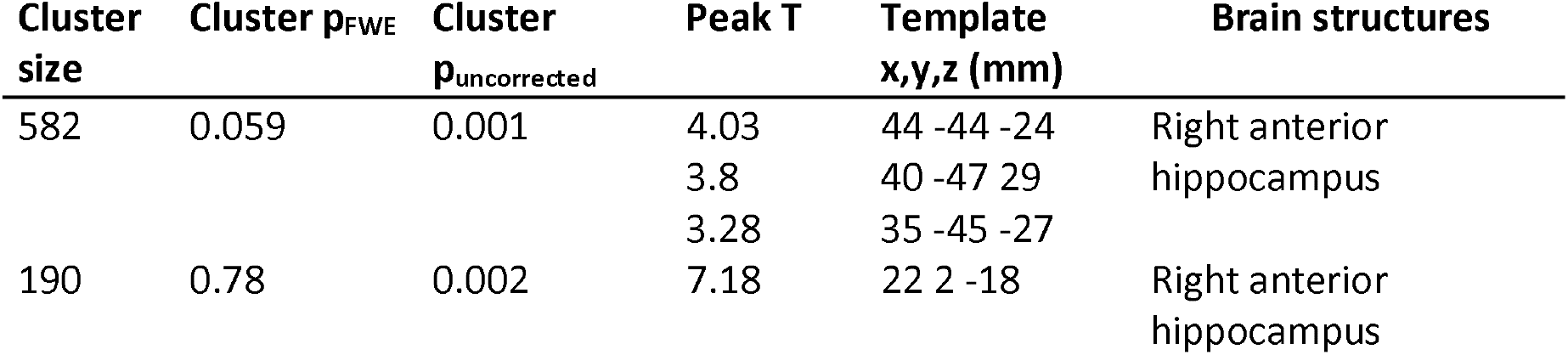
In a subset enriched with CSF data, SN MRI contrast correlates with novelty activation in 2 clusters in the right anterior hippocampu

**Table S3:** ANOVA F score results for diagnostic group differences for all relevant CSF and cognitive measures

| dependent variable | Statistics |
| --- | --- |
| **CSF measures** |  |
| total tau | F(3,67)=14.81 p<0.0001, N=71 |
| Aβ42/40 | |
| ratio | F(3,67)=11.87 p<0.0001, N=71 |
| phosphotau | F(3,67)=10.81 p<0.0001, N=71 |
| **Cognitive parameters** |  |
| recognition memory | F(3,156)=19.5 p<0.001, N=160 |
| Global cognition | F(3,156)=59.73 p<0.0001, N=160 |
| Visual factor | F(3,156)=28.24 p<0.0001, N=160 |
| Working memory Factor | F(3,156)=31.14 p<0.0001, N=160 |
| Memory Factor | F(3,156)=85.06 p<0.0001, N=160 |
| Language Factor | F(3,156)=69.56 p<0.0001, N=160 |

## Notes

### Competing Interest Statement

The authors have declared no competing interest.

### Funding Statement

The study was funded by the German Center for Neurodegenerative Diseases (Deutsches Zentrum fuer Neurodegenerative Erkrankungen; DZNE), reference number BN012. F.K is supported by the German Federal Ministry of Education and Research (BMBF, funding code 01ED2102B) under the aegis of the EU Joint Programme: Neurodegenerative Disease Research (JPND). He is paid from a stipend awarded by the medical faculty in Magdeburg.
E.D. and M.B. are supported by Deutsche Forschungsgemeinschaft (DFG, German Research Foundation) Project-ID 425899996 SFB 1436

### Author Declarations

Our dataset is a subset from the multicenter DELCODE study (Jessen et al., 2018). The Institutional Review Boards of all participating study centers of the DZNE approved the study. All participants gave written informed consent before inclusion in the study. DELCODE is retrospectively registered at the German Clinical Trials Register (DRKS00007966) (04/05/2015).

